# Unlocking the transformative potential of data science in improving maternal, newborn and child health in Africa: A scoping review protocol

**DOI:** 10.1101/2024.07.31.24311286

**Authors:** Akuze Joseph, Bancy Ngatia, Samson Yohannes Amare, Phillip Wanduru, Grieven P. Otieno, Rornald Muhumuza Kananura, Fati Kirakoya-Samadoulougou, Agbessi Amouzou, Abiy Seifu Estifanos, Eric O. Ohuma

**Author notes:** **Corresponding author:** Akuze Joseph; London School of Hygiene & Tropical Medicine, UK.

## Abstract

**Introduction:** Application of data science in Maternal, Newborn, and Child Health (MNCH) across Africa is variable with limited documentation. Despite efforts to reduce preventable MNCH morbidity and mortality, progress remains slow. Accurate data is crucial for holding countries accountable, tracking progress towards realisation of SDG3 targets on MNCH, and guiding interventions. Data science can improve data availability, quality, healthcare provision, and decision-making for MNCH programs. We aim to map and synthesise use cases of data science in MNCH across Africa.

**Methods and Analysis:** We will develop a conceptual framework encompassing seven domains: Infrastructure and Systemic Challenges, Data Acquisition, Data Quality, Governance, Regulatory Dynamics and Policy, Technological Innovations and Digital Health, Capacity Development, Human Capital, Collaborative and Strategic Frameworks, data analysis, visualization, dissemination and Recommendations for Implementation and Scaling.

A scoping review methodology will be used including literature searches in seven databases, grey literature sources and data extraction from the Digital Health Initiatives database. Three reviewers will screen articles and extract data. We will synthesise and present data narratively, and use tables, figures, and maps. Our structured search strategy across academic databases and grey literature sources will find relevant studies on data science in MNCH in Africa.

**Ethics and dissemination:** This scoping review require no formal ethics, because no primary data will be collected. Findings will showcase gaps, opportunities, advances, innovations, implementation, areas needing additional research and propose next steps for integration of data science in MNCH programs in Africa. The findings’ implications will be examined in relation to possible methods for enhancing data science in MNCH settings, such as community, and clinical settings, monitoring and evaluation. This study will illuminate data science applications in addressing MNCH issues and provide a holistic view of areas where gaps exist and where there are opportunities to leverage and tap into what already exists. The work will be relevant for stakeholders, policymakers, and researchers in the MNCH field to inform planning. Findings will be disseminated through peer-reviewed journals, conferences, policy briefs, blogs, and social media platforms in Africa.

**ARTICLE SUMMARY:** *Strengths and limitations of this study:*

➣ This scoping review is the first to examine the role and potential of data science applications in maternal, newborn and child health (MNCH) in Africa, with assessments on healthcare infrastructure, data quality improvement, innovative data collection and analyses, policy formulation, data-driven interventions, technologies for healthcare delivery, and capacity building.
➣ We will conduct systematic searches across multiple databases (PubMed, Scopus, Web of Science, Google Scholar, CINAHL, EMBASE, and Ovid) and grey literature.
➣ Focusing on studies that have used data science we will synthesise our findings with detailed explanations, informative charts, graphs, and tables.
➣ The study will deliver actionable recommendations for stakeholders engaged in MNCH policy formulation, strategic planning, academia, funders and donors, and clinicians aimed at improving MNCH outcomes in Africa.
➣ Our scoping review will primarily rely on published literature in English, therefore, will omit valuable insights that may have been published for non-anglophone and francophone regions of Africa.

## BACKGROUND

The 2015 Sustainable Development Goals (SDGs) require countries to commit to ending preventable maternal, newborn and child deaths by 2030. These goals include reducing under-five mortality in each country to at least 25 per 1000 live births, at least 12 per 1,000 live births for neonatal mortality, and globally at least 70 per 100,000 live births for maternal mortality (1, 2). There needs to be high-quality data on all pertinent indicators in order for nations to be held responsible for their promises (3). Even though countries have made significant work towards achieving these targets, progress is still sluggish, particularly when it comes to birth and deaths registration, and survival and well-being of the most vulnerable (4). UNICEF monitors child well-being through 48 indicators (5) for example, birth registration; education; protection from harm; clean and safe surroundings; and poverty eradication in life, but data gaps remain a significant challenge in many countries (6)

Similarly, over 190 countries worldwide offer data to different United Nations agencies on various indicators like mortality, stillbirth, morbidity, preterm birth, and low birthweight, among other variables (7). However, it is remarkable that the world’s regions with the highest burdens, like Africa, faces a Data Rich Information Poor (DRIP) syndrome(8-10). Accurate data on maternal newborn and child health (MNCH) morbidity and mortality are essential for holding leadership in African countries accountable and achieving the goals of reducing preventable deaths and improved wellbeing (11). In addition to accountability, accurate data would guide better-targeted interventions for the most vulnerable to obtain the most significant impact despite limited resources. Countries need to set priority agendas for MNCH, there is a critical need for data-driven approaches to identify and prioritize key areas. (12-18). During the recent Coronavirus pandemic, there were new advancements using data science in tracking and reporting global mortality such as rapid mortality surveillance and epidemic response within the World Health Organisation’s (WHO)’s Toolkit for Routine Health Information Systems Data (19). However, the pandemic also significantly disrupted MNCH indicators tracking, as well as essential services like immunisation and broader health care provision, including widespread school closures (5, 20-23).

Data science, an interdisciplinary field amalgamating scientific methodologies, algorithms, and systems, is instrumental in deriving insights and knowledge from structured and unstructured data (24). It synthesises elements from mathematics, statistics, computer science, and specific domain knowledge such as MNCH to facilitate data engineering, organisation and management, analysis, and interpretation and dissemination of large volumes of data for informed decision-making (25). Over the last two decades, data science has been applied in several sectors, including healthcare, finance, and marketing, to solve complex challenges, forecast outcomes, and inform strategic approaches (26). Data science applications have also enabled tracking and progress of several SDG targets with improvements in interoperability and improved routine information systems (27, 28). In MNCH, especially in Africa, various applications have been documented, for example predicting infant mortality in Rwanda using ML (29), Prediction of poliovirus, vaccine coverage and immunisation programs in Africa (30, 31) and using data science approaches for decision-making and utilisation of maternal healthcare services in rural Ethiopia (32, 33). With data science applications, there is potential to integrate African MNCH data with other planetary datasets like temperatures, seasonality, rainfall, air pollution, flooding, and NASA’s space data (24).

Data science initiatives elsewhere (Europe, the US, and Asian settings) have been found useful in improving data quality, care and service provision, planning and priority setting as well as implementing targeted interventions for MNCH for example digitisation of health information systems (34-37). Utilising data science in MNCH, mortality and morbidity tracking, data collection, and reporting has the potential to improve the quality, comprehensiveness, and utilisation of data in Africa. Fortunately, many efforts to harness data science in Africa are underway such as District Health Information Software 2 (DHIS2), digital payments, mobile money, ehealth and mhealth platforms (38). However, many of these efforts act in isolation and are not very well coordinated and are not implemented as part of national efforts that can be used to guide policy decisions.

In this study, we conduct a scoping review that aims to review the use, role and contribution of data science to improved MNCH in Africa in recent years. Specifically, the study aims to provide a landscape of data science in MNCH by mapping where, how, and for what MNCH outcomes data science approaches have been applied to in the African region.

### Study rationale

The World Health Organisation has developed a five-year global strategy for digital health (39). It has also established a digital health atlas (https://digitalhealthatlas.org/) to track planning, development, implementation and scale-up of digital health initiatives. On the Digital Health Atlas, projects at different stages of development and their potential impact are shown. Within Africa over 600 initiatives exist to date, however, only 316 focus on the MNCH thematic area and have little information about their stages of implementation (38). Therefore, the extent to which data science has been used to improve MNCH is largely unknown for most of these digital health initiatives. Based on the results of the scoping review, we will also discuss the existing gaps and opportunities for data science in MNCH, illustrate the future trajectory of this emerging field in Africa, and highlight why it is a key pillar for accelerating progress toward national and regional targets. The findings of our review will fortify the knowledge base on data science in MNCH, stimulate further research into this field, as well as highlight case studies and impact made in different African countries. This will also provide a valuable reference for decision-makers to identify potential areas for investment, policy intervention, and collaboration in the field.

### Study objectives

To map and synthesise use cases of data science applications in maternal, newborn, and child health in Africa. We will also highlight case studies and identify existing gaps and opportunities and next steps for further improvement and integration of data science in MNCH in Africa.

## METHODS AND ANALYSIS

### Conceptual model

This section presents the proposed conceptual approach which will be adapted for data extraction and synthesis.

### Conceptualising data science review for Maternal Newborn and Child Health in the African Context

In our scoping review, we shall use a conceptual approach to structure the investigation of how data science has been applied in the field of MNCH in Africa notably at clinical, community and district or regional levels. Our conceptual framework (Figure 1 and Box 1) draws inspiration from the Measure Evaluation’s Health Information System Strengthening (HISS) model designed to assess, plan and improve health information systems in low-and middle-income countries (LMICs) (40). Our adapted model goes beyond the HISS model by incorporating additional domains specific to data science (notably technological innovations, capacity development for data science skills, policy and regulatory considerations and recommendations for implementing and scaling). This approach shall define the scoping review into interconnected domains that shall cover existing gaps, opportunities, and strategic imperatives for integrating data science into MNCH initiatives across the continent. The conceptual framework is designed to guide a comprehensive exploration of the current state, identify gaps and barriers, and spotlight opportunities for harnessing data science in the MNCH context more effectively in Africa.

**Figure 1:**
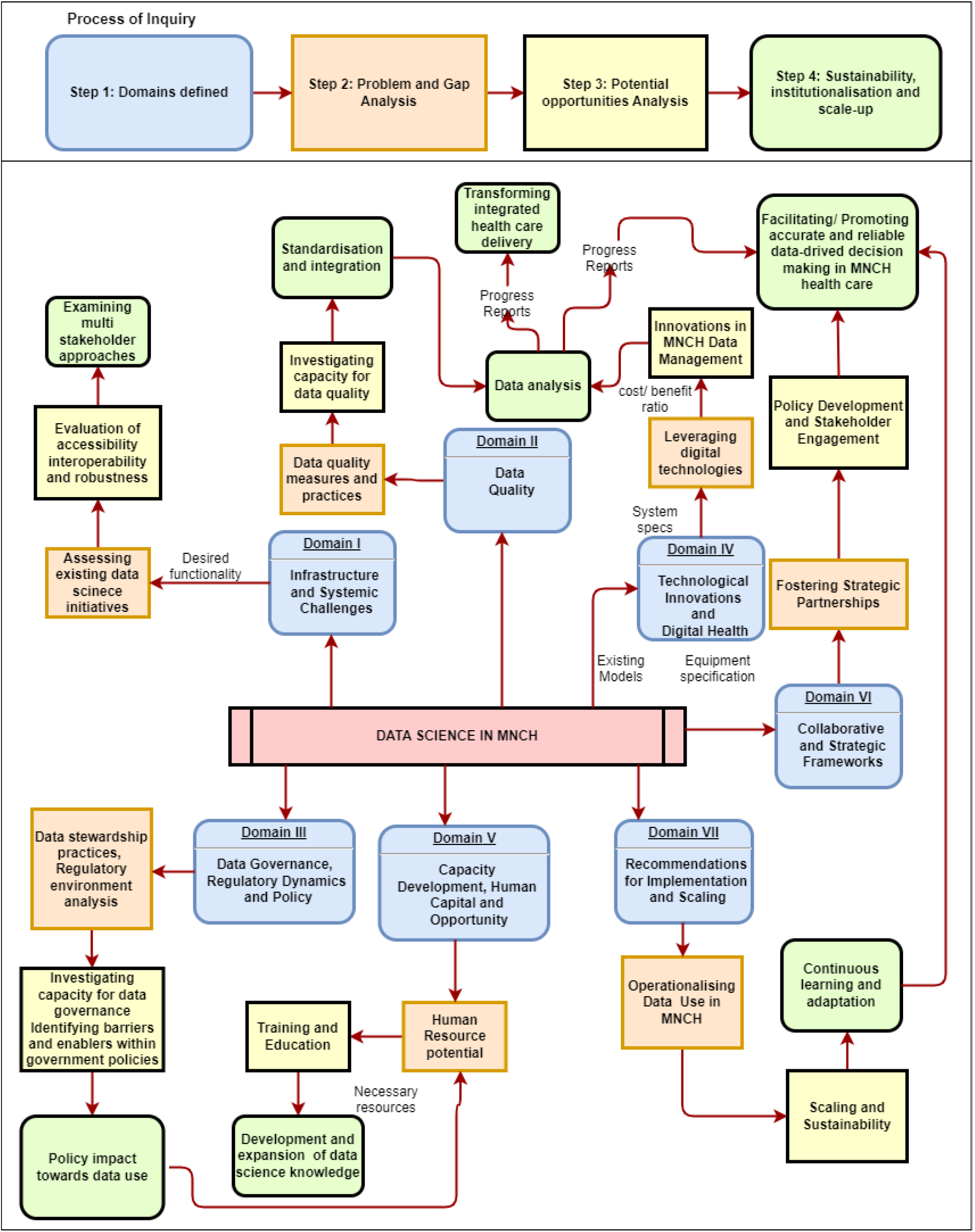
Conceptual framework for harnessing data science to improve MNCH in Africa.

#### Box 1

Conceptual framework for harnessing data science to improve MNCH in Africa

**Domain I. Infrastructure and Systemic Challenges**

Assessing the existing data science initiatives for data infrastructure and data use: evaluation of the accessibility, interoperability, and robustness of current health data systems and data use ecosystem for integrated care and personalised medicine. Examining the multi-stakeholder approaches that impact the utility of MNCH data.

**Domain II. Data Quality**

Standardisation and integration: Addressing the challenges related to data standardisation and quality (accuracy, completeness and reliability of data) to facilitate accurate and reliable health decision-making utilising data science tools and techniques.

Data Analysis for Quality Assurance: Methodologies and frameworks for analysing data quality, to ensure data accuracy, completeness, and reliability.

**Domain III. Data Governance, Regulatory Dynamics and Policy**

Data stewardship practices: Investigating the capacity for effective data governance, focusing on the ethical management and use of health data.

Regulatory environment analysis: Examination of the regulatory landscape affecting the application of data science in MNCH, identifying barriers and enablers within government policies and their impact on moving it forward or hindering innovation.

Compliance and Ethical Guidelines: Ensuring that data science practices comply with local and international regulations and ethical standards.

Policy Impact Assessment: Assessing how changes in policy can either promote or hinder the use of data science in MNCH.

**Domain IV. Technological Innovations and Digital Health**

Leveraging digital technologies: Exploring the potential of mobile health, telemedicine, and Artificial Intelligence to revolutionise MNCH data collection, analysis (including advanced data analytics), and application.

Innovation in MNCH Data Management: Assessing the impact of data science tools and methodologies in transforming integrated health care delivery, personalised medicine and MNCH outcomes.

**Domain V. Capacity Development, Human Capital and Opportunity**

Human Resource Potential: Strategies for building data science capabilities for MNCH to maximise the opportunities.

Training and Education: Prioritising the development and expansion of data science knowledge and skills for data analysis among MNCH health professionals and implementing partners

**Domain VI. Collaborative and Strategic Frameworks**

Fostering Strategic Partnerships: Identifying and leveraging partnerships and collaborations across sectors to advance data science applications in MNCH. Engaging stakeholders to promote data-driven decision-making in MNCH health care.

**Domain VII. Recommendations for Implementation and Scaling**

Operationalising Data Use in Health: Developing actionable strategies for the implementation of data science initiatives in maternal, newborn, and child health care. The role of data analysis in operationalising data use in MNCH for data-driven decision making

Scaling and Sustainability: Recommendations for scaling successful interventions and ensuring their sustainability through continuous learning and adaptation.

### Scoping review protocol design

We propose to use a scoping review design guided by the methodological framework developed by Arksey H and O’Malley L (41), because the implementation of data science within MNCH in Africa at regional, national, and sub-national levels is an emerging field with poor documentation currently. The Arksey H and O’Malley L framework proposes four stages to identify, collate, and summarise the existing literature with scientific rigour, these stages include:

Stage 1: Identifying research question

Stage 2: Identifying relevant studies and data sources

Stage 3: Selection

Stage 4: Collating, summarising of data and synthesis

The scoping review shall springboard our synthesis of the seven domains listed in our conceptual framework. We shall present the screening process using the Preferred Reporting Items for Systematic Reviews and Meta-analyses extension for scoping review (PRISMA-ScR) flow diagram (42).

#### Stage 1: Identifying research question

We have five research questions that we intend to answer notably:

1) Where have the data science projects and initiatives in MNCH been implemented in Africa?
2) What are the experiences and practices of data science in MNCH within Africa?
2) What are the existing gaps and opportunities in using data science in MNCH for Africa?
3) How can we leverage the identified gaps and opportunities to advance data science in MNCH for Africa?
4) What is the future trajectory of data science in MNCH in Africa?
5) What are the opportunities and challenges for effective use and scale-up of data science to optimise decision-making and improve MNCH outcomes in Africa?

#### Stage 2: Identifying relevant studies and data sources

##### Searching strategy

A comprehensive literature search will be conducted in the following databases: PubMed, Scopus, Web of Science, Google Scholar, CINAHL, EMBASE, and Ovid (Table 1 and Supplementary Table S1). In addition, reference lists and peer-reviewed studies will be searched. Other sources of unpublished work, such as websites and blogs by both organisations and individuals, will also be searched through searching for relevant reports obtained from within our existing networks, relevant organisations and government policies published before December 2023. All global multi-country and African data from the WHO’s Digital Health Initiatives Atlas shall be downloaded and reviewed with focus on projects implemented with components of the MNCH thematic area.

**Table 1:** Summary of proposed search terms.

| No. | Proposed search components and key words* |
| --- | --- |
| 1 | Data science AND maternal health OR newborn health OR neonatal health OR child health OR perinatal health AND Africa |
| 2 | Data science AND maternal health OR newborn health OR neonatal health OR child health OR perinatal health AND country (where country represents one of each country in Africa) |

\*Detailed search terms by database are in the appendix table S1

##### Inclusion and Exclusion criteria

Articles and any relevant literature published in English and discussing the status and progress of data science approaches in MNCH in Africa. We will exclude articles without a specific focus on Africa as well as those not highlighting the application of data science in MNCH and systematic and scoping reviews.

#### Stage 3: Selection of articles

##### Document screening and selection

The screening and selection process will include: removal of duplicates, review of the titles to exclude any irrelevant articles (peer-reviewed and grey literature), then a review of the executive summaries or abstract to further exclude any other documents and finally a review of the entire reports for the remaining documents. Two reviewers, one for peer-reviewed articled and one for grey literature will review the articles. A third reviewer shall review the data from the Digital Health Initiatives Atlas to identify projects in MNCH thematic area.

##### Data extraction and collection

Upon reaching an agreement on the final documents for the scoping review, a data extraction process will be undertaken by one reviewer who will enter the data into Rayyan for data management (43). Due to limitations of scope, time and budgetary, we shall use only one reviewer for this. The data extraction process will focus on obtaining information about the author of the document, year, organisation, population, country, donor, study aims and objectives, study design or methods and approaches used (including sample size), context, intervention (type, duration, recipients), key concepts, comparisons made, and main outcomes. The extracted data will further be collated, synthesised, and analysed. For the Digital Health Initiatives Atlas data, similar attributes will be extracted in addition to the project name, software, and challenges.

#### Stage 4: Collating, summarising of data and synthesis

This process will involve reviewing the data extracted and summarised using descriptive analysis. The purpose of this is to synthesise and present an overview of all articles reviewed. Where possible, maps, graphs and tables will be used to visualise the different attributes of data science use in MNCH in Africa basing on our proposed conceptual framework.

### Patient and public involvement

Since this is a protocol for a scoping review, patients and the public were not involved in the design or proposed research.

### Proposed timeline

Conceptualising of the scoping review including the framework began in December 2022. From January 2023 to December 2023, we began drafting a concept note with consultations with African researchers to develop concepts on “The State of Data Science in Africa” on other thematic areas of global health (notably environmental and planetary health, value for data, infectious diseases, policy etc). These efforts are coordinated by African Population and Health Research Centre (APHRC), and National Institute of Health (NIH) Fogarty International Center and have been funded using pooled resources from NIH, Wellcome Trust and Bill and Melinda Gates Foundation. We also formulated the African MNCH Data Science writing team for the scoping review. Data extraction begin in June 2024, and we anticipate that the scoping review will be complete by the four quarter of 2024.

## ETHICS AND DISSEMINATION

The scoping review of already published literature on data science in Maternal, Newborn, and Child Health (MNCH) within African contexts shall not require Ethical approval, because only published and publicly accessible data and literature shall be used.

For dissemination, the findings of this scoping review aim to be shared broadly with stakeholders in the MNCH thematic area, including healthcare professionals, policymakers, and academic researchers. The strategy includes publication in peer-reviewed journals, presentations at conferences related to health informatics and maternal health, and summaries for non-academic audiences to ensure wide accessibility. Additionally, the review will leverage digital platforms and social media to enhance reach and engagement, facilitating the translation of research findings into practical interventions that can be implemented to improve MNCH care and outcomes in Africa. This approach ensures that the insights derived from the application of data science in MNCH are disseminated effectively, fostering collaboration and innovation in the field. We shall also disseminate findings through Nature Africa, policy briefs, blogs, webinar series and workshops.

## Discussion

This study shall contribute to a publication series on the Status of Data Science in Africa by APHRC and NIH showcasing the expected transformations and advances in fields of biomedical and behavioural research with data science applications (44). Data Science is an emerging field especially on the African continent, therefore, the review on its applications in the MNCH context is timely and covers and addresses issues around the whole African continent. One limitation of the study is that only publications in English shall be included, therefore limiting our understanding of the application and impact of Data Science in MNCH from other non-anglophone or francophone countries in Africa.

## Supporting information

Supplementary Material Table S1: Proposed detailed search terms

## Data availability

No underlying data is associated with this article

## Extended data

Open Science Framework (OSF): Unlocking the transformative potential of data science in improving maternal, newborn and child health in Africa: A scoping review protocol https://doi.org/10.17605/OSF.IO/8PYNC

## Twitter/X

Akuze Joseph @JosephAkuze

Philip Wanduru @phillwdru

Rornald Muhumuza Kananura @KananuraRornald

Bancy Ngatia @bnbancy

Samson Yohannes @zigniter

Grieven Otieno @GrievenOtieno

Fati Kirakoya @FKirakoya

Abiy Seifu Estifanos @AbiSe

Eric O. Ohuma @ohumaeric

## Author Contributions

EOO conceptualised the idea, AJ drafted the manuscript in coordination with EOO. AJN, EOO, and BN developed the research questions and study methods and contributed meaningfully to the drafting and editing; PW, RMK and EOO reviewed and contributed to the manuscript developed; PW, FKS, AA, RMK, SY and ASE aided in developing the research question and study methods, contributed meaningfully to the drafting and editing, and approved the final manuscript. All authors reviewed and approved the final version of the article for publication.

## Acknowledgements

We thank the National Institute for Health (NIH) Fogarty Division of International Science Policy Planning and Evaluation, African Population and Health Research Centre (APHRC), and other partners for forming “The State of Data Science for Health in Africa writing initiative”, coordinating the writing process and funding the writing workshops and meetings held in Nairobi that our authors participated in. We also thank the Data Science for Africa-Initiative Co-chairs (Catherine Kyobutungi, Emile Chimusa and Kofi A. Amegah) for their leadership and guidance of the consortium.

## Data statement

The data is publicly available and a table of the articles that will be included in the review will be provided as a summary table.

## Patient consent for publication

Not required

## Provenance and peer-review

Not commission; externally peer reviewed

## Supplementary material

The authors provided the supplementary material

## Open access

This article is available under the terms of the Creative Commons Attribution 4.0 International (CC BY 4.0) License. This license allows for the copying, distribution, modification, and building upon the work, for any use, as long as proper credit is given to the original author, a link to the license is included, and it is indicated if modifications were made to the original work. More information can be found at: https://creativecommons.org/licenses/by/4.0/

