## Supplementary Material Table S1: Proposed detailed search terms for "Unlocking the transformative potential of data science in improving maternal, newborn and child health in Africa: A scoping review protocol"

**Table S1: Proposed detailed search terms**

| **Geography** | **Methods** | **Thematic areas** |
| --- | --- | --- |
| "Africa" OR "Algeria" OR "Angola" OR "Benin" OR "Botswana" OR "Burkina Faso" OR "Burundi" OR "Cameroon" OR "Cape Verde" OR "Central African Republic" OR "Chad" OR "Democratic Republic of Congo" OR "Republic of Congo" OR "Cote d'Ivoire" OR "Djibouti" OR "Egypt" OR "Equatorial Guinea" OR "Eritrea" OR "Ethiopia" OR "Gabon" OR "Gambia" OR "Ghana" OR "Guinea" OR "Guinea Bissau" OR "Kenya" OR "Lesotho" OR "Liberia" OR "Libya" OR "Madagascar" OR "Malawi" OR "Mali" OR "Mauritania" OR "Mauritius" OR "Morocco" OR "Mozambique" OR "Namibia" OR "Niger" )) OR ALL=(( "Nigeria" OR "Reunion" OR "Rwanda" OR "Sao Tome and Principe" OR "Senegal" OR "Seychelles" OR "Sierra Leone" OR "Somalia" OR "South Africa" OR "South Sudan" OR "Sudan" OR "Swaziland" OR "Tanzania" OR "Togo" OR "Tunisia" OR "Uganda" OR "Zambia" OR "Zimbabwe" OR "Southern Africa" OR "East Africa" OR "Central Africa" OR "Northern Africa" OR "West Africa" | "data science" OR "artificial intelligence" OR AI OR "machine learning" | “maternal” OR “neonatal” OR “child” OR “perinatal” |
